# Willingness to work in assisted living facilities: The Ghanaian nurses perspectives

**DOI:** 10.1101/2020.07.01.20144238

**Authors:** Irene Korkoi Aboh

## Abstract

**Objective:** This study explored nurses willingness to work in assisted living institutions for the aged.

**Background:** Assisted living institutions are multifunctional facilities that provide clinical and ambulatory (day hospital) care for somatic and psychogeriatric elderly with multiple pathology, disability, and handicaps.

**Methods:** The study used a mixed-methods approach in which qualitative data was collected first before the quantitative data. Data was collected through focused group discussions (FGDs) and questionnaires from 248 respondents with age ranging from 20 to 58 years from October 2016 to January 2017 with 8 missing from the quantitative data. Four pertinent questions were sked both in the study. Sampling was convenient and purposive from 4 different health institutions in the metropolis. Data from the FGDs were digitally recorded and transcribed verbatim. Quantitative data was entered in SPSS version 23 and cleaned. Both sets of data were coded and analysed.

**Result:** The nurses appreciate the increase in the number of the aged in their communities; they think community members prepare towards their ageing by using their children as security, and the idea of assisted living was enthusiastically supported, but with the proviso that it would need to be ran by a private entrepreneur. Almost all the 240 respondents said that government should establish an institution for the aged and they would be willing to work in such an institution, if only it will run by a private entrepreneur.

**Conclusion:** The nurses also think that families are now becoming more nuclear; that is why caring for the aged has become a problem, thus creating a need for assisted living facilities.

## Introduction

Caring for older people is a humbling experience, regarded as unselfish and loving assistance given to another (Eriksson, 2010). It is about responsibility and watching over the other and can be carried out by a lay person (Israelsson-Skogsberg & Lindal, 2017). Assumptions are that nursing practice focuses on what matters for nurses caring for the elderly, thus listening attentively to what the elderly says about themselves and their lives, gives cues on what is meaningful for them and how their quality of life, peace of mind, body and soul is (Bernick, 2004). The challenges of caring for older individuals can be a scary task for health care providers because it requires multi-specialty expertise with the focus of meeting the individual’s psychological, cultural, spiritualty and physiologic needs (Goldstien & Anapolsky, 2004).

Policies in Western societies have been geared towards enhancing institutions as a home domain for residents through regulation of sheltered accommodation and nursing care facilities for older persons and increasing each person’s autonomy (Clancy & Mahler, 2016). From a caring perspective, there is an emphasis on the staff and the organisation respecting the older person’s integrity and personal boundaries, both relational and corporeal (Rostgaard & Szebehely, 2012). Well-being in an older person’s everyday life is dependent on respect for their integrity and dignity which is very important (Galvin & Todres, 2013).

## Background

Nurses practitioners (NPs) provide direct patient care and it involves cognitive assessment, reviews, ordering and /or reviewing tests and medication, and liaising with family members (Bentley, Minstrell, Bucher, Spoule, Robinson & Stirling, (2015). Referrals by NPs include referrals to geriatricians, to general practitioners, to allied health professionals (e.g. physiotherapists, dieticians), and to social service (e.g. home care and dementia support programmes) (Schadewaldt, McInnes, Hiller & Gardner, (2013). The role of a registered nurse in the residential aged care facility is multifaceted and includes miscellaneous forms of caring, managing the individual impact of ageing, the opportunities of clients and their respective regarding care needs (Drew, 2011). They are required to provide leadership and guidance in care directives, which includes training learning and development of other subordinate staff and team members, and assisting clients make informed decisions, particularly in relation to treatment choice, palliative pathways and end-of-life issues (Caruana, 2008). It is also noted that RNs are the clinical leaders in aged care in effective management of staff retention within their service (Fussell, McInerney & Patterson, 2009).

Among the various staff types in a geriatric centre are contract staff and aids including certified nursing assistants, home health aides, personal care aides or assistants, and medication technicians or aides. Social workers include licensed social workers, graduates with a bachelor’s or master’s degree in social work in adult day service centres and residential care communities, and medical social workers (Harris-Kojetin & Sengupta, 2013, p16). The normal working hours per day in nursing homes and residential care settings for adult day services, home health agencies and hospices have been reported as averaging 8 hours per day in three 24-hours section shifts (National Association for Home Care and Hospice & Hospital and Healthcare Compensation Service (NAHC/HH/HCS), 2009), with a worker-to-caregiver ratio of 3:26 (MetLife Mature Market Institute (MMMI), 2010).

Nurse practitioners spend considerably more time with the elderly than do general practitioners attached to these facilities. They are more accessible, able to initiate more timely care, visit elderly people in their homes and thereby increase access to care for those who are not mobile or not able to drive themselves to service (Australian Government Department of Health (AGDH), 2016). NPs undertake more comprehensive assessments of older persons than other registered nurses; this means that there is better quality of clinical information to be used by the broader healthcare team in ordering diagnostic tests or initiating appropriate medicines, and timelier treatment schedule for acute facility and can circumvent complications of the condition (AGDH, 2016). From an economic perspective, nurse practitioners in Australian aged care save dollars for government funders through their timely and accurate care interventions. Economic efficiencies were gained through reductions in: unnecessary transfers to acute health facilities, ambulance costs, hospital bed days, and thus hospital cost (Davey et al., 2015).

Demand for home-based care is rapidly increasing as baby boomers are ageing and advanced medical technologies are extending the life expectancy of disabled and chronic patients, with an estimate that by 2050, 27million people will need some type of long-term care (Department of Health and Human Services (DHHS), 2003). The National Home Health Aide Survey (NHHAS) also estimated that almost 1.46million older people were receiving care in 2007 and 7.2 million had received care and were discharged in 2000 (Kim & Antonopoulos, 2011). The authors added that over 14,000 agencies were in the business of recruiting and training caregivers for serving clients.

Assisted living is any residential setting not licensed as a nursing home but which provides or arranges personal care and routine nursing in a homelike residential setting (Lewin-VHI, 1996). It is regulated according to its state or region, which also reflects different philosophies about who should be served in these residential long-term care settings and the relationship envisaged between assisted living and nursing homes (Mollica, 2000). These facilities, are living arrangements that provide personal care and health services for people who may need assistance with activities of daily living but wish to live independently. The level of care provided is not as extensive as that which may be provided in a nursing homes (Genworth, 2014). Assisting living is not an alternative to a nursing home, but an intermediate level of long-term care. One facility may look like a modern high-rise apartment building; another might look like a quiet suburban town home community. Others may resemble a resort hotel or a country club, but there are some generalizations (SeniorAdvisor.com, 2017). The homelike nature of the settings also varies a great deal among and within state, with some assisted living settings providing single-occupancy apartments and others providing shared accommodations in board-and-care settings with two or more persons per room (Frytak & Kane et al., 2000).

Apartment-style assisted living, by design, offers privacy and the opportunity for autonomy. It also exposes residents to risks of everyday life associated with cooking and bathing and tends to afford staff less opportunity for protective surveillance. Whether the service is provided by internal staff, outside home care agencies, or a combination of these, assisted living tends to be more lightly staffed than nursing homes (Frytak & Kane et al., 2000).

## Materials and methods

### Design and setting

A mixed-method study design and a sequential procedure in which the researcher sought to elaborate on or expand the findings of one method with another method. The study began with a qualitative investigation for exploratory purposes, followed by a quantitative investigation with a large sample so that there could be generalization of results to the population (Creswell, 2003, p16). Therefore, qualitative data was collected first and questionnaires were designed to answer questions that were missed during the qualitative phase.

The study was carried out in the Cape Coast Metropolitan Assembly, one of the 17 districts of the Central Region of south Ghana. The Metropolis is further divided into 25 sub-districts for administrative purposes. Of the 25 sub-districts, 10 were randomly selected using SPSS version 20 software, and an adjoining community was used to test study instruments. These 10 sub-districts were further classified into three different zones: urban (elite residential communities), peri-urban (either urban or rural, but densely populated) and rural (lacking almost all social amenities). Subsequently, the zones were demarcated as zones A, B, and C. All the government health facilities in the Central region are in the metropolis. Four health facilities were used for the study, considering one teaching hospital, a Metropolitan hospital, a quasi-hospital, and a psychiatric hospital, in which all the categories of nurses were present. Looking at the individual strength of these health facilities, a proportional-to-size approach was used in allocating sampling to distribute populations from which participants were recruited. It was concluded by the principal investigator (PI) and research team that the Regional Hospital, which was a referral point for all major and complex cases, would be allocated 40% of estimated sample size, because all categories of nurses was seen there.

This was followed by 30% for the psychiatric hospital which served the middle belt of the country and one of the three government psychiatric hospitals. The district hospital and the University hospital, which is a quasi-hospital, followed with 20% and 10% respectively.

### Sampling and data collection techniques

A total of 248 nurses was originally proposed for the study so 248 questionnaires were sent out, but only 240 questionnaires were collected because these were the respondents willing to give the needed time and information due to their busy schedules. Forty were recruited to participate in the focus group discussions (FGDs). The inclusion criterion was all categories and ranks of nurses, but excluding midwives on the grounds that these were in a specialized branch of nursing that did not have much contact with the aged.

The PI and the research assistant collected all the qualitative data. An unsaturated interview guide was used to conduct discussions. The four main questions for all the groups were as follows; (a) What was the welfare of the aged in regard to activities of daily living? (b) How did the aged prepare for their current situation? (c) On a scale of 1 to 10, how strong were they? (d) Would they be willing to accept responsibility in caring for the aged in an assisted living environment? The questionnaire included both structured and semi-structured questions, which were divided into five sections: Section A –personal information; Section B – preparedness of aged for their ageing; Section C – current care practices looking at activities of daily living; Section D – care that needed to be added to what the aged presently received: Section E – willingness of nurses accept responsibility in caring for the aged in an assisted living environment.

An introductory letter and a gatekeeper from University of KwaZulu-Natal and the Dodowa Research center was sent to the individual facility’s research office and one field worker each was allocated to assist in data collection. Appointments were scheduled for commencement of data collection for both the focus group discussions and the questionnaires. The facility research officer (FRO) sensitized the nurses in preparation for the focus group discussions and a period of seven days was allocated to organized participants for the focus group. After each discussion, a day was scheduled for those who had attended the discussion to meet with other nurses for responding to the questionnaires so that key informants would send the message to other parts of the facility through grapevine communication about the impending data collection. The age range of target population was 20 to 58 years. Strengths Weaknesses Opportunities Threats (SWOT) analysis was used to explain the reason for the research to participants and respondents. Five field workers and a research assistant were recruited, based on their knowledge and experience of the topic under study and were recruited, based on their knowledge and experience of the topic under study and were given three days of training. Data collection began in October 2016 and ended in January 2017. Although three FGDs were scheduled, difficulties due to nurses’ workloads and distance of the from the facilities could not allow for 3 but 2 FGDs, followed by the collection of the quantitative data. The discussions lasted between 40 minutes and one hour.

A total of 240 questionnaires were completely answered. A quota system was used where each health facility was given a number of questionnaires to answer through the research officer with the help of the field workers. Any nurses met at the data collection point was included and taken through the questionnaire. Those who were busy were given a day or two to complete and return the form, with follow-ups from field workers. Participants and respondents were assured of confidentially and anonymity before each procedure began by giving them consent forms to sign, and those who were not ready to be part of the study were given the opportunity to withdraw.

### Data management and analysis

The recorded FGDs were transcribed verbatim from native language into English by the PI and a trained research assistant. The transcripts were checked for accuracy and quality and cleaned for anonymity by the PI. When no discrepancies were identified, the files were coded for analysis. The methods of analysis were interpretive descriptive analysis to gain insight into the willingness of nurses to accept responsibility in caring for the aged in an institution (Richie & Lewis, 2003).

In an initial step, the content of the data files was read to identified the major distinctive themes that provided meaningful constructs that illuminated the concept under study. Three key elements of interpretative descriptive analysis were followed: *detection* (which involved identification), assigning the substantive content and *displaying* dimensions of the topics under study. Six major themes and three sub-themes emerged: trends of ageing, preparedness of the aged for ageing, government plans for the aged, caring and caregivers, assisted care (sub-themes: *cultural ideologies, inception and funding*), and acceptance of responsibility.

## Results

### Participant characteristics

The nurses showed mixed feelings about the intent of the study, but were willing to give researchers the needed information. Of the 40 participants recruited for the FGDs, all had successfully completed their basic nursing education; six had also completed their baccalaureate, and four had master’s degree. They were all Christians and married. Three were females and 37 males; ages ranged from 25 to 44 years. Some had children while others did not. The demographic characteristics from data collected from the questionnaire blended with the characteristics of those in the FGDs.

About 48.8% of eligible respondents in the quantitative study were in the 20-29 years age group, and just 2.9% were in the 50-59 years age group. Majority of those who had time for questionnaire were female, single and Christians. More than half (53.8%) had a diploma qualification as their highest level of educational attainment, and they were either general or psychiatry nurses with the rank of staff nurse.

### Trends in ageing

Ageing and death is inevitable, when participants were asked if they had noticed that the population of the aged had increased in their communities, they thought it was due to the aged being pushed into the hospitals because of festive occasions and because community members did not want the aged to be in their way. Some knew about this trend in research reports form dissemination of results to the hospital; others had not because they spent most of their time in the hospitals and scarcely interacted with the community. Another participant agreed that the aged were increasing in number possible because of public health strategies or health promotion activities. All the participants echoed the point made by one contributor that,

> *Yes … we have seen that because the aged are being pushed into the hospitals on festive occasions…. We the young guys when we invite our friends to our homes, we do not want them to see our frail parents so we hide them*. ***FM4***.

From information in a report disseminated to the hospital, a participant added that

> *We have noticed that the longevity of Ghanaians has increased through a survey. And Ghanaians are growing older and our population is now between 25-30million*. ***FB1***.

Another participant added that

> *I can also say that due to good and proper medical care, the young ones are growing hence the increment in population*. ***FM3***.

Table 3 shows awareness of the increasing number of aged in our communities. About 66.6% said ‘Yes’ to the question and 33.4% said ‘No’. The table also shows the responses on whether the aged were kept in the hospitals on festive occasions and about 67.9% responded ‘Yes’.

**Table 1.** Research sites and activities.

| Site | Zones | Activity/Status of Facility/<br>Number of participants | Activity/Number of respondents |
| --- | --- | --- | --- |
| Site 1 | A | Regional Hospital –Level C category of hospital | Questionnaires – 96 |
| Site 2 |  |  |  |
| Site 3 |  |  |  |
| Site 4 |  | Metropolitan Hospital Level B | Questionnaires -- 48 |
|  |  | category of hospital |  |
|  |  | FGD-20 ---FM |  |
| Site 5 | B |  |  |
| Site 6 |  | * | * |
| Site 7 |  | Quasi – Hospital – Level B | Questionnaires – 24 |
| Site 8 |  | category of hospital |  |
| Site 9 | C |  | Questionnaires --72 |
| Site 10 |  | Psychiatric Hospital Level C |  |
|  |  | category hospital |  |
|  |  | FGD – 20 ---FP |  |

\* No institution is located in this zone –no activity was carried out there.

**Table 2.**
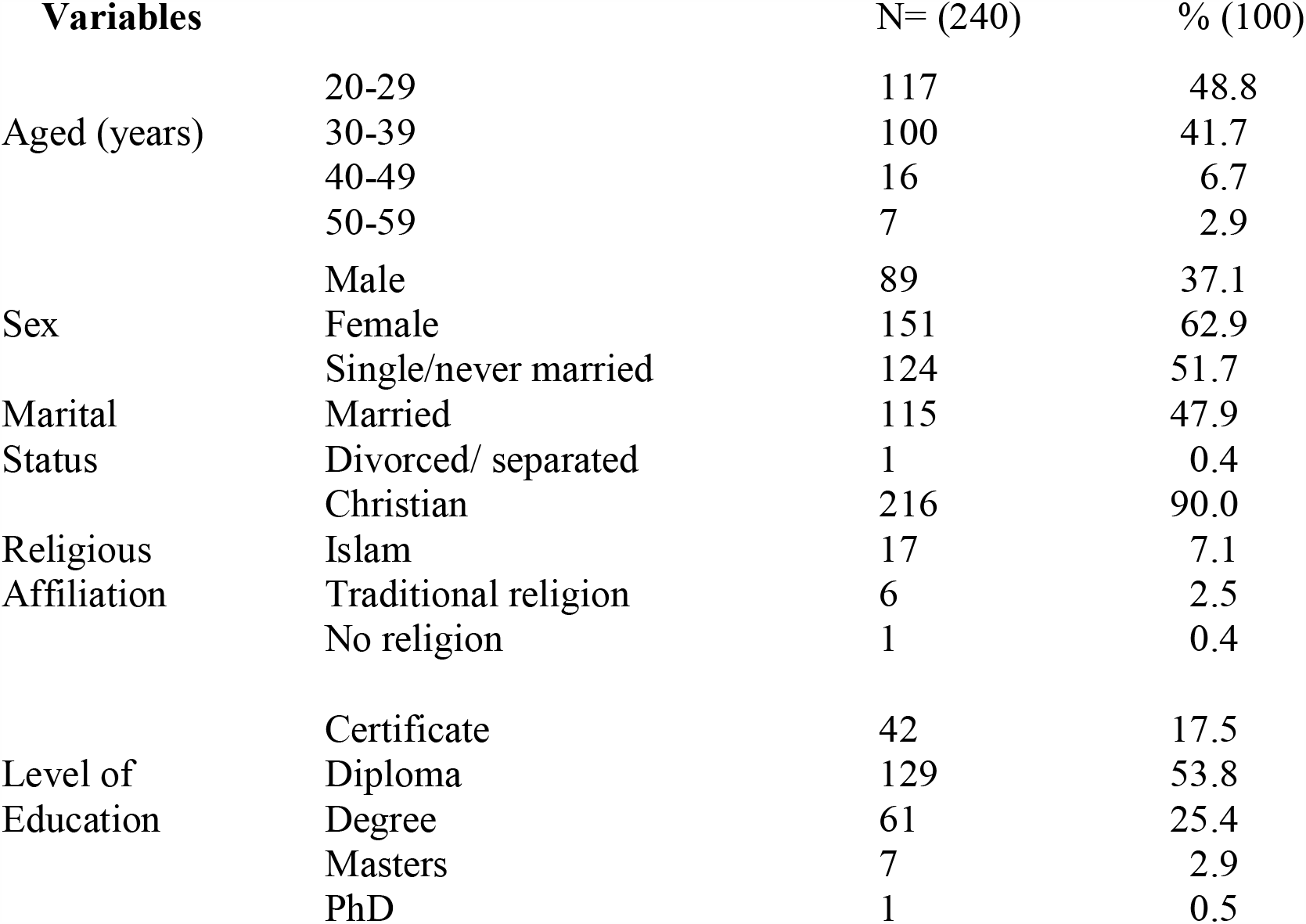
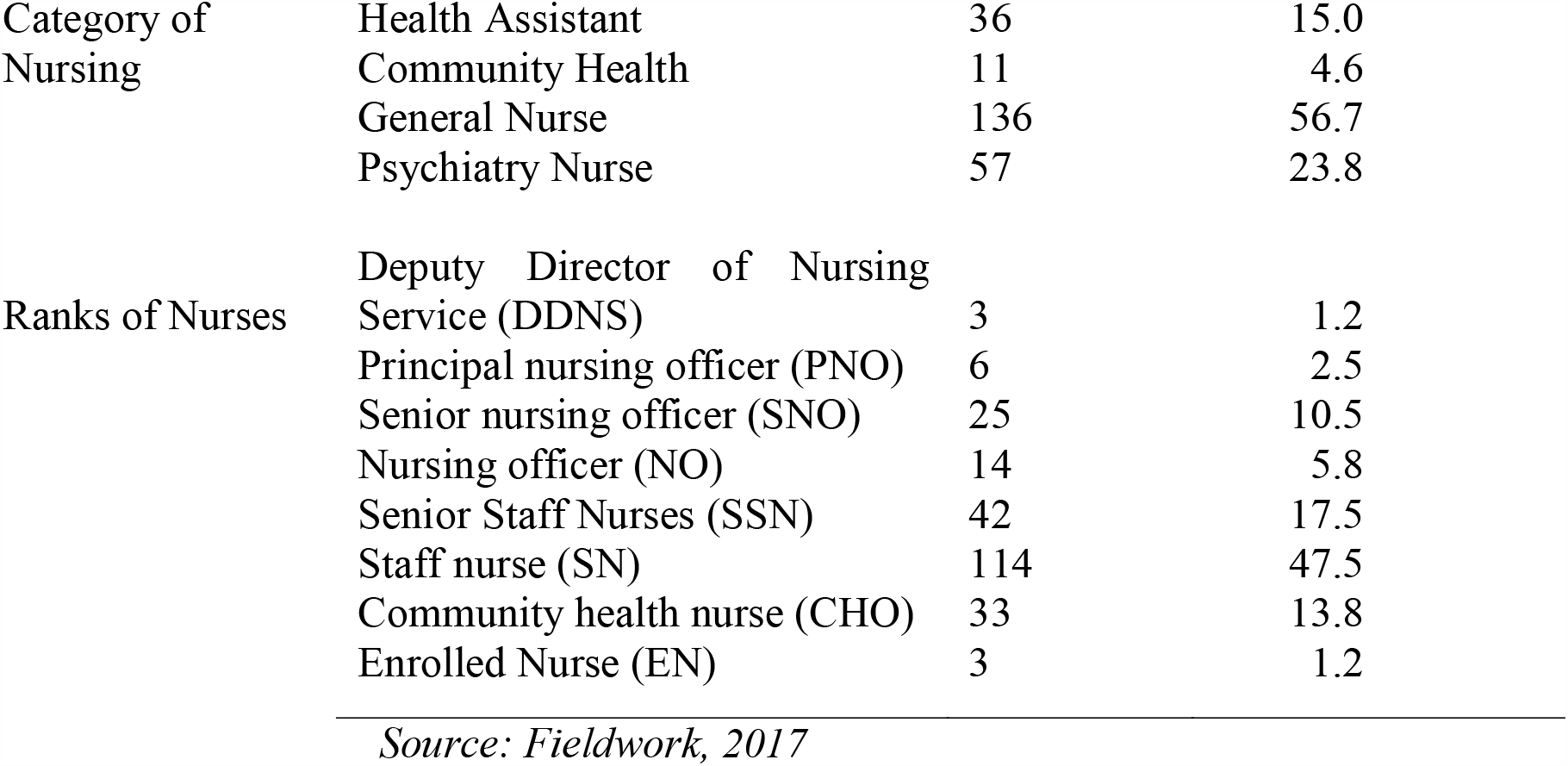
Quantitative data on demographic characteristics of nurses.

**Table 3.** Quantitative data on Nurses’ awareness of issues of the aged.

| Response | Yes (%) | No (%) |
| --- | --- | --- |
| Awareness of increasing number of aged | 160(66.6) | 80(33.4) |
| Aged being sent to the hospital | 163(67.9) | 77(32.1) |
| **Community ways of securing for the future** |  |  |
| Variable | N | Percentage |
| Having many children | 45 | 18.8 |
| Looking after other people's children | 31 | 12.4 |
| Applying for person | 25 | 10.4 |
| Nothing that I know off | 110 | 45.8 |
| Others | 29 | 12 |
| **Government plans for the aged** |  |  |
| Variable | N | Percentage |
| Cared for by the children | 11 | 4.6 |
| Housed in their homes | 14 | 5.8 |
| Look for caregivers for them | 12 | 5.0 |
| Nothing so far | 203 | 84.6 |
| **Relationship between caregiver and aged** |  |  |
| Parents | 65 | 27.1 |
| Nephew/niece | 135 | 56.2 |
| No relation | 33 | 13.8 |
| Tenant | 5 | 2.1 |
| Living alone | 1 | 0.4 |
| Own sibling | 1 | 0.4 |

*Source: Fieldwork, 2017*

### Preparedness for ageing

When participants were asked how the community planned for their future, they all responded that people planned through ensuring a certain amount of security by taking good care of their children for reciprocation of care in the future.

> *Our society plan through their kids … they try to take care of their kids very well or any family member living with them, so that in their old age they can also rely on them for care*. ***FM6***

Others said they knew people who planned by building houses so that they could retire and come back home to their loved ones before dying. Other participants suggested that those who plan do it without the knowledge of their family while others do not plan at all:

> W*hat I see is people think of residence, where they will spend the rest of their lives when they are old … a place to live after pension. Some do it secretly without their family’s knowledge … others too plan their lives secretly by saving toward their funeral and buying of caskets*. ***FM8***.

Another participant added that,

> *for those who travel to seek greener pastures … they make sure they build houses so that when they come back … they will get a place to stay … because they believe that it is good to die around your loved ones*. ***FM6***.

One participant thought that only the educated plan for their future,

> *some of our people invest into businesses so that when they go home, they can be getting dividend from their investments and these are the educated ones. Apart from these nothing*. ***FM3***.

Tables 3 again shows perceived community ways of securing the future. 45.8% had no idea how the community members plan for their ageing, followed by 24.4% who said others look after people’s children so that care will be reciprocated in future and18.8% said some people have their children as their future security.

### Government plans for the aged

Participants were asked what government did for the aged; they all seemed to know about the national health insurance scheme (NHIS), the pension scheme, LEAP, and the EBAN because the programmes are ongoing and they had a fair idea of them:

> *the NHIS … it is a policy that is free for people 65 ears and over, they enjoy free premium by the policy’* ***FM2;***
>
> *to me this (pension) plan is limited to the literate i*.*e. those who have ever had formal classroom education. … It is based on their active years’ contribution … they are given something (money) until they die;* ***FM4***.

Those who had heard of LEAP were not sure how it really worked:

> *they are given some money (LEAP) especially those who are too poor to cater for themselves’* ***FM4;*** *the only one l know of … is the LEAP but I do not know how it works, I heard they go and register for it’*. ***FP1***.

One participant thought only the disabled were eligible for the plan and he was corrected.

> *I thought that programme was for disables*. ***FM7***

There was general agreement that ‘the aged have been added’, and one participant commented that’

> *For my area the LEAP is only for widows who are old … or who are above 65 years and more but for the male aged I have not heard anything’*, ***FM6***.

Another added that he had heard of the EBAN:’

> *I also have heard that the aged are given priority whenever they go to the health facilities for attention or the EBAN for the state owned or metropolitan buses’* ***FM6***.

Table 3 indicates responses on government planning for the aged, a majority (84.6%) said that government had no plan so far; 5.8% said the government plan was for the aged to be housed in their own homes and 4.6% said the government plan was for aged to be cared for by children irrespective of the environment.

### Caring and caregivers

When asked about the care and caregivers of the aged they know in their communities, the participants all agreed that although the caregivers of the aged they know in their communities, the participants all agreed that although the caregivers think they are doing their best in caring for the aged it was not good enough. Some suggested that the root of the problem of poor caring practices was that the extended family system was breaking down and everybody was now busy, so the aged were left unattended. One participant said he had often seen old people in the company of young grandchildren about eight years old looking for directions to a destination and ending up in a different part of the town. It was report that

> *Caring is not good because we are moving to the nuclear family system that is why we are facing these challenges. In the nuclear family setting, the aged are left alone at home …… so they need more caregivers … But with the extended family system their children and their kids to their grandmothers so they tend to enjoy the interaction of these grandchildren. Meanwhile in our setting because of our extended family system there was always someone who is self-employed and ready to care for the aged*. ***FP1***

Another participant added that

> *In our communities, women are more and they suffer the neglect than the men so the government in its own wisdom targeted the women*. ***FM6***

Another described how a friend’s grandmother was poorly treated by his own mother,

> *People do neglect the aged …. And they leave them to their fate … because at their age they are branded as witches and then neglected. Just yesterday a friend called me and complained about how his mother was maltreating his grandma … she hardly cooks for her, so when he questioned the old lady on how she feeds … she said she sleeps on empty stomach when there is no food … she eats when there is food and stays put that is how she lives and I think it is very much appalling*. ***FM2***

Caring is traditionally done by females in the indigenous African home; participants were quick to add that it was appropriate for the female child to look after the aged in the community even through caring could also be done by a male child. Indicative comments were:

> *Caring is the responsibility of the daughter*. ***FM4***
>
> *From where I come from the aged stays with either the first born or the last born irrespective of the sex*. ***FM6***
>
> *In my community, it is the responsibility of the last born to care for their aged but if the aged has a daughter then the daughter will leave wherever she is and go and care for her parents … she must come and stay by running a routine between her aged parents and her nuclear family*. ***FM3***

Another participant reported that grandchildren were made to stay with their grandparents and he was an example,

> *I stay with my grandmother; because of the low educational status of the community, they do not plan for ageing. What they do is that they rely on their husbands if they are alive then it is the responsibility of their husbands to take care of their house and everybody. … So, when there is a girl in the family … they will let the girl come and stay with her grandparents and help with their little things that they will need. I went to stay with my grandmother so I did have chores because my sister had left for school … but now another girl is there*. ***FM7***

Table 3 shows the relationship between caregivers and the aged they take care off. A majority (56.2) were close family members of the second generation, and 13.8% of caregivers were not blood relatives of the aged in their care. One aged person was reported to be living alone and another with a sibling.

### Assisted living

Assisted living illustrates the shift from caring as a humanitarian service to caring as a business in the broader health care arena in the 1970s where it has become a consumer-driven industry that offers a wide range of options, levels of care, and diversity of services (Lockhart, 2009). When asked what they thought about assisted living, participants asked what kind of nursing care entailed. The PI took the opportunity to explain the concept of assisted living to them using the SWOT approach. There was some confusion among participants as they deliberated among themselves and the PI gave them more time to discuss their fears and uncertainties. There were expressions of disapproval as to why people would want to give up their cultural role of caring for their own to be replaced by something, they had no idea of. Other participants felt that government should assign nurses to take care of the aged in their homes so that they could also attend to spiritual needs. Another group was happy because they knew people who were looking for such places for their aged. There were also participants who were convinced that day care would work but felt that there should first be suitable policy for the programme before its inception.

### Cultural ideologies

Those opposed to the idea of assisted living thought that government should come out with a policy on care for the aged that would be binding on every household in the country.

> *I have my reservations about this ‘assisted living …. You see, a lot of people will be running away from their responsibilities. …. I suggest there should be a policy or law before this programme comes into being … so that everybody would be made to take care of their aged … and penalties accorded if those rules are breached or flouted … if people neglect their parents, grandparents and it known … these culprits would be punished … people who do not want to look after their aged could dump them there*. ***FM7***
>
> *I am not in support with this idea because when we were young, they took care of us. …. We were defecating and urinating on them and doing all sort of things on them so it is now time for us to also care for them. Now, when it is time for reciprocation of care we are trying to dodge our responsibility; leaving it for someone else to do. … As my brother rightly said people will shed their responsibility to other people. … My suggestion is if education will be effective on how to take care of the aged. I think it will be good*. ***FM8***

### Programme inception

There were various comments on the possible inception of the programme participants:

> *I saw this nursing in a foreign movie once … and it was very fine. …. Due to our cultural beliefs, the old ones believe there is a spirit of the environment so they would not like to leave their environment. …. But if government too can do that for us it will be fine. …. Nurses can take nursing to their door step so that it will be like door to door nursing care. … Caring for the spiritual needs of the aged is also very important so if they can train nurses, it will be very fine. …*.. *Every month or twice a month a nurse can go and visit them at home and involve family members in their care*. ***FM6***
>
> *Left to me, government must train more community health officers (CHOs) to handle the aged just like they are handling the children. The old people also believe that they are the custodians of the land … so if you ask her to go and stay somewhere … it will not be proper. The uneducated would want their parents to stay in the house. Meanwhile the young ones cannot say they will not work to look after their aged parents … so the government can train nurses solely for this programme to go around the homes to help with caring for the aged*. ***FM5***
>
> *In Ghana now, we are becoming enlightened so there should be education for people to be aware of these innovations. It should be in and out in the institution and home setting. We must push this thing forward; there will be ups and downs of the programme while it is taking root. When it takes off … I believe all these challenges we are bringing up will be taken care off so long as we pay attention to them*. ***FP8***
>
> *The day care will be proper; our communities have been structured such that every community in Ghana has a community or social centre … so the chief should agree to let his people to be trained … so that they take off the aged at the community centre … day-care type will be more feasible. … the day care will work; … it will be a place where they will have a lot of fun then they come home in the evening. …. This will work in Ghana*. ***FP3***
>
> *I also think the churches can help … because in my church they are trying to use the social centre to be doing something creative for the aged. So, that the aged who can walk will come and play indoor games when they are bored at home then go back at the end of the day. …. Some of the tithes we pay at church is used to do this activity*. ***FP1***

## Funding

Participants offered extended comments on how the programme should be funded:

> *As part of our pension scheme, we must be advised so that the same thing to be done on our aged institutions … we have complaints from the community about people neglecting their aged*. ***FP5***
>
> *Government can do it through private partnership so that it will be regulated like the health insurance system. This could be one aspect of the health sector, to be the other aspect that would make life complete*. ***FP4***
>
> *Government should go into partnership with private entrepreneurs so that they can handle this programme as a business entity*. ***FP3***
>
> *Ehmm! We are being advised to secure other means of pension plans as workers in our active life; …. So if we are saying that government should take up responsibility it can also take up that part of setting up policies to care for the aged … Just like it did for the 3tier system of the existing pension plan … so that such funds could easily be accessed. … More so the emphasis should be on social support … We are worried about people in the communities complaining about neglecting their aged parents … but I also want to say that anybody who is sensitive enough to identify that their aged parents need care in such a home or secured services in a home or looking for someone to cater for them is supportive enough … Ahhaaa!!! So, if we should promote this practice rather than undermine it from the scratch we could go a long way*. ***FP8***
>
> *As for Ghanaians, we will agree to pay for the something … we start and then we let it hang an example is the mental hospital saga … People will bring their wards with promises … but when they go home that is all. … They do not do as they promised and it will become a burden for the government. We do not want such things to repeat itself because people would be dodging their responsibilities and start blaming the government when things go wrong, … however there should be another way of reaching out to the aged through government’s point of view. They should be in their own home environment for government reach out to them through LEAP*. ***FM6***

### Acceptance of responsibility

Questions that were missed from the FGD question guide for participants were captured in the questionnaire for respondents. Question on willingness to work in an institution was captured and answered by 240 respondents.

In the descriptive statistical analysis, the variables score showed a statistically significant p-value of 0.043, showing there is an association between respondents agreeing to the establishment of assisted living and readiness to work in an assisted living facility. Table 4 also displays the response of respondents to the establishment of, and working in an aged care institution or an assisted living environment of, and willingness to work in an aged care institution or an assisted living environment; while 2.1% were not in favour of the idea.

**Table 4.**
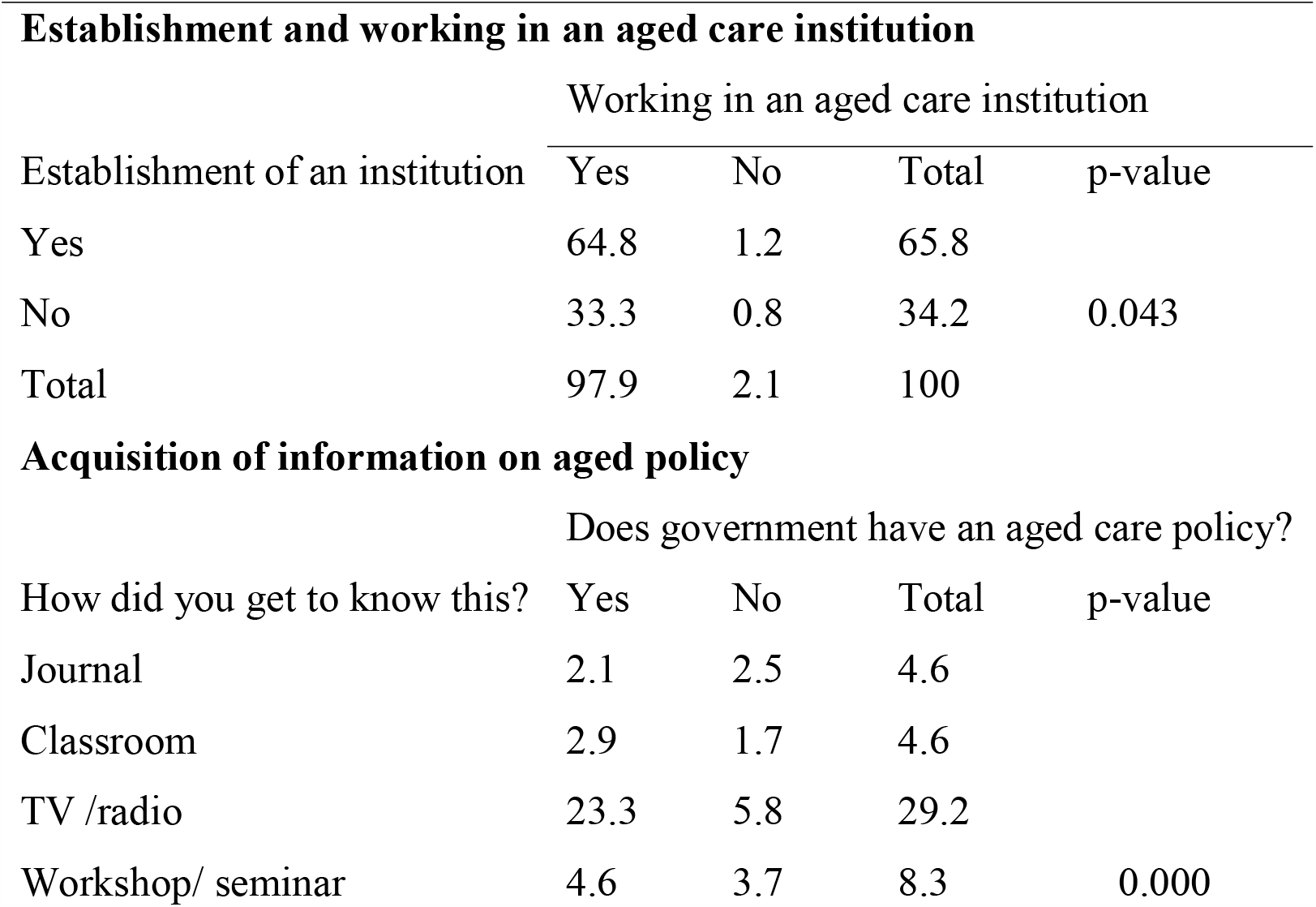
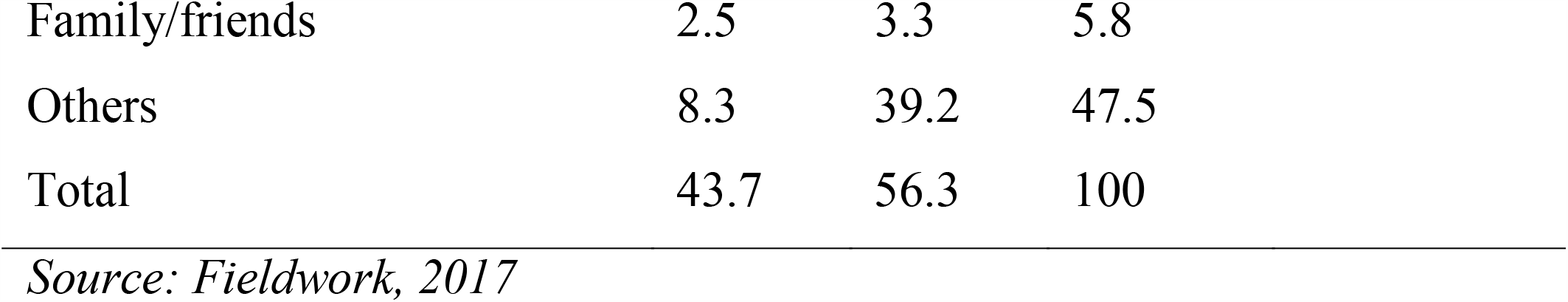
Quantitative Data on Nurses Interest in Aged Issues.

| **Establishment and working in an aged care institution** |  |  |  |  |
| --- | --- | --- | --- | --- |
|  | Working in an aged care institution |  |  |  |
| Establishment of an institution | Yes | No | Total | p-value |
| Yes | 64.8 | 1.2 | 65.8 |  |
| No | 33.3 | 0.8 | 34.2 | 0.043 |
| Total | 97.9 | 2.1 | 100 |  |
| **Acquisition of information on aged policy** |  |  |  |  |
|  | Does government have an aged care policy? |  |  |  |
| How did you get to know this? | Yes | No | Total | p-value |
| Journal | 2.1 | 2.5 | 4.6 |  |
| Classroom | 2.9 | 1.7 | 4.6 |  |
| TV /radio | 23.3 | 5.8 | 29.2 |  |
| Workshop/ seminar | 4.6 | 3.7 | 8.3 | 0.000 |

|  |  |  |  |
| --- | --- | --- | --- |
| Family/friends | 2.5 | 3.3 | 5.8 |
| Others | 8.3 | 39.2 | 47.5 |
| Total | 43.7 | 56.3 | 100 |

*Source: Fieldwork, 2017*

In the descriptive statistics, there was statistically significance association between having knowledge of government policy and source of this knowledge with a p-value of 0.000. Table 4 indicate acquisition of information on the policy for the aged and how this information was acquired. A significant number of participants (39.2%) had no idea of any policy on aged care, even though they were getting information about the aged from, in most cases, two or more sources. Less than a quarter of the participants said that, ‘Yes’, they knew about this policy from TV and radio.

## Discussion

This study set out to determine the willingness of skilled nurses to work in an aged care institution or assisted living environment. More than 90% agreed to the inception of the programme and to accepting responsibility to work in that environment. Although participants in the focus group were skeptical about feasibility of the programme, since family members had an obligation to take care of their own even when resources might be scarce, they nonetheless agreed that care for the aged was not good, due to the breakdown of the extended family system and its replacement by nuclear families. The nurses also acknowledged that health promotion strategies had increased the longevity of Ghanaians. Appreciation was expressed for a documentary on changing family structures, showing that as people live longer and have fewer children, family structures are transformed, leaving older people with fewer options for care (Dobrainsky, Suzman & Hodes, 2007). Population ageing is a phenomenon that results from declines in fertility as well as increases in longevity: two trends that are usually associated with social and economic development (POPFACTS, 2015).

Participants and respondents did not know how community members plan for t heir ageing. Some said that caring for one’s children and other people’s children was used as a guarantee for the future in the form of reciprocated care when one becomes frail. According to Karlberg (2008). Karlberg cited an Akan proverb to illustrate this: “If someone looks after you to grow your teeth, you must also look after him to lose his”. This system has been a security for people without children, however, in a modernized society this system is difficult to maintain. Ultimately the system fails if these children move away to find work. Van de Geest 2002, in a study in central Ghana, found that growing doubt around family solidarity meant that loyalty shifted from lineage to conjugal family, and that middle-class, urbanized Akan families had begun to keep non-nuclear relatives at bay, avoiding claims by the *abusua* on their income and possessions.

Lack of awareness of government plans could be due to the nurses being too fully occupied with workplace commitments to get involved in community issues or communications on policy implementation from other sectors. All respondents knew about the NHIS, which had a direct bearing on their job descriptions, as suggested by the Abuja Declaration which called for African heads of state to allocate at least 15% of annual government budgets to the health sector. In addition, it was expected that this financial support would increase substantially over decades (Carrin et al., 2008).

Assisted living will be a new caring practice in the Ghanaian health sector if accepted in the country, with study participants finding the concept difficult to understand until it was explained to them. As word got around about the concept in the course of the sequential data collection in the study, almost all respondents (98%) showed interest in the establishment of an aged care institution and in working in such an institution or accepting deployment to one by their employers.

In terms of the technology acceptance model (as applied in the field of information technology and computing), which uses a simplified interpretation of beliefs that affect technology acceptance (CHIR, 2017), there are two kinds of belief: perceived ease of use refers to the belief that using a particular system would be free of physical and mental effort, and perceived usefulness. Perceived ease of use refers to the belief that using a particular system would be free of physical and mental effort, and perceived usefulness is the degree to which an individual believes that using a particular system would enhance his or her job performance. These distinctions been applied to the adoption of technology in workplaces, such as health care facilities (Holden & Karsh, 2010), in studies focusing on physicians (Pare, Sicotte, & Jacques, 2008), nurses (Tung, Chang & Chou, 2008), and other clinicians (Schaper & Pervan, 2007) as adopters of the new technology. Similar distinctions may help to explain the dilemma faced by nurses in this study as to ease of use of assisted living facilities, coupled with goal-driven concerns about whether output in their nursing care for the aged will enhance quality of care and improve the status of situation in the community.

The concept of funding was rightly suggested as needing to be of the social security type, where the individual contributes to his/her ageing in line with government-controlled policy. It was also agreed by participants that all programmes controlled by policies are well managed and sustained.

## Conclusion

The purpose of this study was to explore and describe the willingness of nurses to accept responsibility for working in an assisted living facility. Issues that emerged were the existence of poor caring practices that would favour the introduction of assisted living, and the importance for study respondents of perceived usefulness of people in the community, but also thought it would be worthwhile to try the assisted living concept because lives were at stake. There was also valuable input from respondents about the need for formulated policy to control the programme and using entrepreneurship to set it going. Study participants confirmed that there are some individuals in the communities who have no caregivers, and that for those who do, the care is not what they would consider adequate. This suggests that this programme, through its multidisciplinary approach to nurses, caregivers and clients, may be meeting needs in the metropolis.

### Ethical approval

The study was approved by the Humanities and Social Sciences Research Ethics Committee of the University of KwaZulu-Natal (HSS/0608/016D) and the Dodowa Health Research Centre (IRB Ghana Health Services) of Ghana (DHRCIRB/06/06/16). Voluntary participation was accorded with written and signed consent.

## Limitations of the study

Convenient purposive sampling procedures were followed, and scheduling the FGDs to accommodate nurses’ work times, workloads and distance from home was difficult, and although PI and assistant conducted altogether three FGDs, two came out well but the third was disrupted by noise from construction work in a last-minute change of venue, affecting the data. The nurses also complained about the number of pages to be read, and there were a lot of distractions from their clients and patients. Problems also arose in trying to coordinate focus group meetings with changes of shift. We also recognized researcher impact on the study participants.

## Data Availability

Data will be available unpon reseanble requet from author.

## Competing interests

The author declares that there is no competing interest with anybody.

## Authors’ contributions

IKA conceived the study, and was the principal investigator to the research. This study is a sub-study of her PhD thesis.

## Author information

IKA is a nurse Educator and lecturer at the School of Nursing and Midwifery, University of Cape Coast, Ghana.

## Acknowledgements

I thank the health facilities that granted permission to conduct this research. Field work was supported by the bursary from the College of Health Sciences, University of KwaZulu-Natal.

## Notes

### Competing Interest Statement

The authors have declared no competing interest.

### Funding Statement

There was no funding

### Author Declarations

The study was approved by the Humanities and Social Sciences Research Ethics Committee of the University of KwaZulu-Natal (HSS/0608/016D) and the Dodowa Health Research Centre (IRB Ghana Health Services) of Ghana (DHRCIRB/06/06/16).

